# Monocyte-endothelial interactions as a targetable node in clonal hematopoiesis-mediated cardiovascular disease

**DOI:** 10.1101/2024.02.20.24303046

**Authors:** Alyssa C. Parker, J. Brett Heimlich, Joseph C. Van Amburg, Yash Pershad, David A. Ong, Nicole A. Mickels, Laventa M. Obare, Ketan J. Hoey, Hannah K. Giannini, Ayesha Ahmad, Caitlyn Vlasschaert, Tarak N. Nandi, Ravi K. Madduri, Samuel S. Bailin, John R. Koethe, Celestine N. Wanjalla, Alexander G. Bick

## Abstract

**Background:** Clonal hematopoiesis of indeterminate potential (CHIP) increases risk of cardiovascular disease yet the molecular mechanisms driving this association remain incompletely understood. We hypothesized that aberrant monocyte-endothelial interactions contribute to CHIP-mediated cardiovascular disease.

**Methods:** We performed single-cell RNA sequencing on blood and peripheral vascular tissue from 4 individuals with TET2 CHIP, 6 individuals with DNMT3A CHIP, and 25 controls. We predicted interactions between monocytes and endothelial cells based on expression of ligand-receptor pairs then modeled interactions between monocytes carrying CHIP mutations with endothelial cells *in vitro*. We performed an *in silico* genomewide perturbation screen to identify genetic targets capable of mediating these interactions and then experimentally evaluated the effect of inhibiting predicted targets on monocyte-endothelial interactions *in vitro*.

**Results:** Expression of ligand-receptor pairs on monocytes and endothelial cells from patients with and without CHIP highlighted differences in signaling likelihood for 6 key ligand-receptor pairs related to transendothelial migration. Co-culture of monocytes with human aortic endothelial cells demonstrated that monocytes carrying CHIP mutations have decreased velocity compared to monocytes without CHIP mutations. The perturbation screen suggested 11 druggable genetic targets capable of rescuing TET2 CHIP monocytes. Experimental inhibition of ICAM1 in endothelial cells and inhibition of CXCR2 in monocytes significantly increased the velocity of *TET2*-mutated monocytes over endothelial cells.

**Conclusions:** CHIP mutations alter interactions between monocytes and endothelial cells. Therapeutics targeting CXCR2 and ICAM1 may restore normal interactions between monocytes and endothelial cells among patients with TET2 CHIP.

## Introduction

Clonal hematopoiesis of indeterminate potential (CHIP) is characterized by the acquisition of somatic mutations in hematopoietic stem cells.^1,2^ CHIP has been associated with adverse cardiovascular outcomes, including coronary artery disease, heart failure, and peripheral vascular disease.^3–6^ CHIP increases the risk of cardiovascular disease (CVD) independent of known risk factors like diabetes.^7,8^ Mouse models of CHIP have suggested that hematopoietic cells carrying mutations in *TET2* and *DNMT3A* are sufficient to induce vascular disease.^9,10^ However, the mechanisms by which mutations in blood cells give rise to vascular disease in human patients are incompletely understood.

Human vasculature is challenging to study because most vascular tissue cannot be easily sampled from living patients. However, adipose tissue is a highly vascularized tissue that can be easily sampled from living patients.^11–13^ We simultaneously sampled peripheral blood and adipose tissue from individuals with diabetes, then performed targeted DNA sequencing to determine CHIP status. Using single-cell RNA sequencing and knowledge of ligand-receptor interactions, we predicted the likelihood of interactions between monocytes and endothelial cells, based on CHIP stats. We identified mutation-specific alterations to monocyte-endothelial interactions for patients with CHIP and sought to attenuate these effects with perturbation of genetic targets. Our findings provide new insights into the effects of CHIP on the interaction between monocytes and endothelial cells and suggest therapeutic strategies that may affect vascular disease risk among patients with CHIP.

## Methods

### Peripheral blood mononuclear cell and adipose tissue sample collection

Patient samples for this study were recruited through the HIV, Adipose Tissue Immunology, and Metabolism (HATIM) study (NCT04451980) and the Clonal Hematopoiesis and Inflammation in the VasculaturE (CHIVE) cohort at Vanderbilt University Medical Center. The study was approved by the Vanderbilt University Medical Center Institutional Review Board (identifiers: 210022 and 201583), and all study participants gave informed consent before sample collection.

All study participants had diabetes at the time of sample collection. Participants from the HATIM cohort contributed both peripheral blood and adipose tissue samples, as described in the original publication.^11^ Participants from the CHIVE cohort contributed only peripheral blood samples, as described in the original publication.^14^

### CHIP calling

CHIP status was determined using an established targeted sequencing assay.^15^ Briefly, DNA extraction from whole blood was performed using Qiagen Mini kits Cat #27104, following standard procedures. Sequencing was performed on an Illumina Novaseq 6000 using a custom gene panel designed to target CHIP driver genes with 600x read depth. Putative somatic mutations were identified using Mutect2-GATK. Variants with total low read depth (<100), low variant allele read depth (<3), and/or variant allele fraction below the threshold for CHIP (<2%) were removed.

### Single-cell RNA sequencing of peripheral blood mononuclear cells

Cryopreserved PBMCs were thawed at 37°C then washed with complete RPMI (RPMI (Corning, #<u>10-040-CV</u>) + 10% FBS + 1% PS, cRPMI) to remove the freezing media. An aliquot of 500,000 cells from each sample was plated with phosphate-buffered saline and incubated for 4.5 hours at 37°C and 5% CO2. Cells were then pooled with unique hashtag antibody oligonucleotide-conjugates (HTOs) for 30 minutes (Biolegend, TotalSeq-B). Samples were prepared with a 10X Chromium 3’ library preparation kit (10X Genomics) and immediately run on a 10X Chromium Controller to create scRNAseq libraries. To ensure capture of a sufficient number of myeloid cells for well-powered comparisons, we enriched for CD14+ cells prior to sequencing for a subset of samples. This was done using an EasySep Human Monocyte Isolation Kit (StemCell Technologies, #19059).

Next-generation sequencing was performed on an Illumina Novaseq6000 at Vanderbilt Technologies for Advanced Genomics core and output was processed with the standard CellRanger pipeline from 10X Genomics. Downstream analyses were performed on Terra.bio.

### Single-cell RNA sequencing data processing

Ambient RNA was removed using the R^16^ (v4.4.0) package SoupX^17^ (v1.6.2). Further analyses were primarily performed with the R package Seurat^18^ (v4.4.0). Cells with greater than 25% mitochondrial reads, <800 total transcripts, or <200 genes were removed. Doublets were identified using the R package DoubletFinder^19^ (v2.0.3). Doublets and cells lacking hashtag oligos were removed. Reads from mitochondrial and ribosomal genes were also removed. Gene counts were normalized using the Seurat function NormalizeData(), followed by FindVariableFeatures() with nFeatures = 3000. Counts were scaled with the ScaleData() function, and dimensionality reduction was performed with the functions RunPCA() and RunUMAP() with dims = 1:15. Batch effects were corrected with the R package harmony^20^ (0.1.1) and clustering was performed with the Seurat functions FindNeighbors() and FindClusters(). Annotation of cell types was performed with scType^21^ (v1.0).

### Differential expression of single-cell RNA sequencing data

scRNAseq analysis is complicated by high levels of sparsity. To address this, we used a metacells approach in which transcriptionally similar cells are collapsed into single measurements. Metacell assignments were determined using the Python^22^ (v3.10.11) package Metacell-2^23^ (v0.8.0). Genes that had at least 10 transcripts in 85% of metacells were considered for downstream analysis. Differential expression was calculated with a negative binomial Wald test and Benjamini Hochberg p-value adjustment from the R package DESeq2^24^ (v1.40.2). Age, sex, and BMI were included as covariates.

### Monocyte-endothelial cell signaling prediction

Cell signaling was predicted using the R package CellChat^25^ (v1.6.1) using standard methods. Monocyte-endothelial signaling was determined by classifying classical monocytes and endothelial cells as both source and target cells, stratified by mutation-specific CHIP status. Statistical significance was evaluated with a Wilcoxon signed-rank test.

### Monocyte-endothelial co-culture and movement analysis

We introduced mutations in *TET2, DNMT3A*, or *AAVS1* (control) into hematopoietic stem cells using CRISPR. We differentiated cells towards the monocyte lineage by culturing them in CD34 expansion media, as described above, for 7 days, followed by 7 days in monocyte differentiation media, containing IMDM + 20% FBS + 0.5% P/S and the following recombinant human cytokines (Gibco): SCF (#300-07-10UG) at 25 ng/mL, IL-3 (#200-03-10UG) at 30 ng/mL, FLT3-L (#300-19-10UG) at 30 ng/mL, and M-CSF (300-25-10UG) at 60 ng/mL. Simultaneously, human aortic endothelial cells (Lonza, #CC-2535) were grown to confluency in 12-well glass-bottom plates (MatTek, #P12G-1.5-14-F) pre-treated with fibronectin (Corning, #356008) using Vascular Cell Basal Medium (ATCC, #PCS-100-030), including 20% FBS and EGM-2 SingleQuots (Lonza, #CC-4176). Monocytes were counted and transferred to the 12-well plates containing endothelial cells. Imaging was performed at the Vanderbilt Cell Imaging Shared Resource Core on a Zeiss LSM 880 with the incubator set at 37* with 5% CO2. Images were taken every 5 minutes for 30 minutes.

After imaging, monocyte movement was tracked using TrackMate^26^ through Fiji.^27^ Cell detection was performed with the LoG Detector with an estimated object diameter of 15 microns. Initial thresholding removed objects called with low confidence. No filters were set for mean intensity ch1 or contrast ch1. Tracking was performed with the Simple LAP tracker. Linking max distance was set to 50 microns, gap-closing max distance was set to 50 microns, and gap-closing max frame gap was set to 1. No filters were set for the number of observations in a track. Average velocity was quantified and compared between groups. Statistical significance was calculated with a two-sample T-test.

### In silico perturbation analysis with single-cell foundation model Geneformer-30M

We predicted genetic perturbations likely to shift CHIP monocytes towards a control state using Geneformer, a foundation model trained on scRNAseq data from approximately 30 million cells.^28^ Using classical monocytes from previously published scRNAseq data for patients with high VAF CHIP,^14^ we fine-tuned the pretrained Geneformer-30M model to distinguish monocytes from people with and without TET2 and DNMT3A CHIP. Fine-tuning of Geneformer was accomplished by initializing the model with the pretrained Geneformer weights and adding a final classification-specific transformer layer. The default hyperparameter values for fine-tuning from the Geneformer-30M Hugging Face model were used. We evaluated the accuracy and macro-F1 of the classifier calculated on the basis of a fivefold cross-validation strategy for which training was performed on 80% of the cells and performance was tested on the 20% cells, repeating for five folds. We extracted the penultimate layer of the fine-tuned classifier and visualized these embeddings using UMAP dimensionality reduction. With our fine-tuned classifier that distinguishes between TET2 and control cells (or DNMT3A and control cells), we performed *in silico* perturbations to predict the effect of genetic knockout on cell state. The Geneformer-30M model and fine-tuning scripts are available on HuggingFace to build a classifier (classifier.py) and *in silico* perturbation (in_silico_perturber.py and in_silico_perturber_stats.py).

Genes predicted by both CellChat and Geneformer were prioritized for *in vitro* validation. Inhibitors were selected to target either a receptor expressed on monocytes or a receptor expression on endothelial cells, based on availability of drugs. For experiments involving drug perturbation of monocytes, cells were replated on day 6 in monocyte differentiation media containing 1 uM, 2 uM, 5 uM, or 10 uM of AZD5069 from MCE and cultured for 24 hours. For experiments involving drug perturbation of endothelial cells, drugs were added to endothelial cells at 0.1 nM, 1 nM, 2 nM, 5 nM lifitegrast sodium (SAR1118) from MCE 24 hours before co-culture. Imaging and analysis of monocyte velocity were performed as described above.

### Visualization

Data visualization was completed with the R package ggplot2^29^ (v3.4.4) and with the Python package seaborn^30^ (v0.13.2). Schematics were prepared using BioRender.com.

### Code availability

Code will be made available on https://github.com/bicklab upon publication. Code for Geneformer-30M is available at https://huggingface.co/ctheodoris/Geneformer.

### Data availability

scRNAseq data will be made available on https://cellxgene.cziscience.com/ upon publication.

## Results

We performed single-cell RNA sequencing (scRNAseq) on 19 peripheral blood mononuclear cell (PBMC) samples and 31 adipose tissue samples from patients with diabetes. Blood samples were obtained from 5 patients with DNMT3A CHIP, 3 patients with TET2 CHIP, and 11 controls (**Table 1**) and resulted in a total of 51,254 cells after filtering low quality cells. We obtained an average of 3,482 reads per cell and 1,584 genes per cell. Adipose tissue samples were obtained from 5 patients with DNMT3A CHIP, 3 patients with TET2 CHIP, and 23 controls (**Table 2**), resulting in a total of 93,975 cells after filtering low quality cells. We obtained an average of 2,972 reads per cell and 1,292 genes per cell. We captured cell types at the expected proportions in both PBMC and adipose tissue samples (**Figure 1a, Figure 1b**).

**Table 1.** Demographic and CHIP-specific details for patients who donated peripheral blood samples.

| Patient | Gene | VAF | Variant |
| --- | --- | --- | --- |
| 1 | Control | NA | NA |
| 2 | Control | NA | NA |
| 3 | Control | NA | NA |
| 4 | Control | NA | NA |
| 5 | Control | NA | NA |
| 6 | Control | NA | NA |
| 7 | Control | NA | NA |
| 8 | Control | NA | NA |
| 9 | Control | NA | NA |
| 10 | Control | NA | NA |
| 11 | Control | NA | NA |
| 12 | DNMT3A | 0.02 | NM_022552:exon20:c.2377delT;p.Y793Tfs*9 |
| 13 | DNMT3A | 0.036 | NM_022552:exon23:c.G2645A;p.R882H |
| 14 | DNMT3A | 0.096 | NM_022552:exon17:c.2007dupC;p.I670Hfs*43 |
| 14 | DNMT3A | 0.022 | NM_175629:exon8:c.856-2A>T;NM_153759:exon4:c.289-2A>T;NM_022552:exon8:c.856-2A>T;NM_001320893:exon3:c.400-2A>T |
| 14 | KIT | 0.021 | NM_000222:exon13:c.C1900T;p.R634W |
| 15 | DNMT3A | 0.028 | NM_022552:exon15:c.1775delA:p.Y592Sfs*59 |
| 16 | DNMT3A | 0.13 | NM_022552:exon23:c.G2645A:p.R882H |
| 17 | TET2 | 0.02 | NM_001127208:exon3:c.G2413T;p.G805X |
| 18 | TET2 | 0.03 | NM_001127208:exon9:c.T4120G;p.C1374G |
| 19 | TET2 | 0.51 | NM_001127208:exon3:c.2867_2868insA:p.L957Ifs*15 |
| 19 | SRSF2 | 0.4 | NM_001195427:exon1:c.C284G;p.P95R |

**Table 2.** Demographic and CHIP-specific details for patients who donated adipose tissue samples.

| Patient | Gene | VAF | Variant |
| --- | --- | --- | --- |
| 20 | Control | NA | NA |
| 1 | Control | NA | NA |
| 21 | Control | NA | NA |
| 22 | Control | NA | NA |
| 23 | Control | NA | NA |
| 2 | Control | NA | NA |
| 24 | Control | NA | NA |
| 3 | Control | NA | NA |
| 25 | Control | NA | NA |
| 26 | Control | NA | NA |
| 4 | Control | NA | NA |
| 5 | Control | NA | NA |
| 27 | Control | NA | NA |
| 28 | Control | NA | NA |
| 6 | Control | NA | NA |
| 29 | Control | NA | NA |
| 30 | Control | NA | NA |
| 31 | Control | NA | NA |
| 32 | Control | NA | NA |
| 7 | Control | NA | NA |
| 33 | Control | NA | NA |
| 8 | Control | NA | NA |
| 9 | Control | NA | NA |
| 12 | DNMT3A | 0.02 | NM_022552:exon20:c.2377delT;p.Y793Tfs*9 |
| 13 | DNMT3A | 0.036 | NM_022552:exon23:c.G2645A;p.R882H |
| 14 | DNMT3A | 0.096 | NM_022552:exon17:c.2007dupC;p.I670Hfs*43 |
| 14 | DNMT3A | 0.022 | NM_175629:exon8:c.856-2A>T;NM_153759:exon4:c.289-2A>T;NM_022552:exon8:c.856-2A>T;NM_001320893:exon3:c.400-2A>T |
| 14 | KIT | 0.021 | NM_000222:exon13:c.C1900T;p.R634W |
| 15 | DNMT3A | 0.028 | NM_022552:exon15:c.1775delA;p.Y592Sfs*59 |
| 34 | DNMT3A | 0.026 | NM_022552:exon10:c.T1220C;p.I407T |
| 17 | TET2 | 0.02 | NM_001127208:exon3:c.G2413T;p.G805X |
| 18 | TET2 | 0.03 | NM_001127208:exon9:c.T4120G;p.C1374G |
| 35 | TET2 | 0.023 | NM_001127208:exon6:c.3595-2A>T |

**Figure 1:**
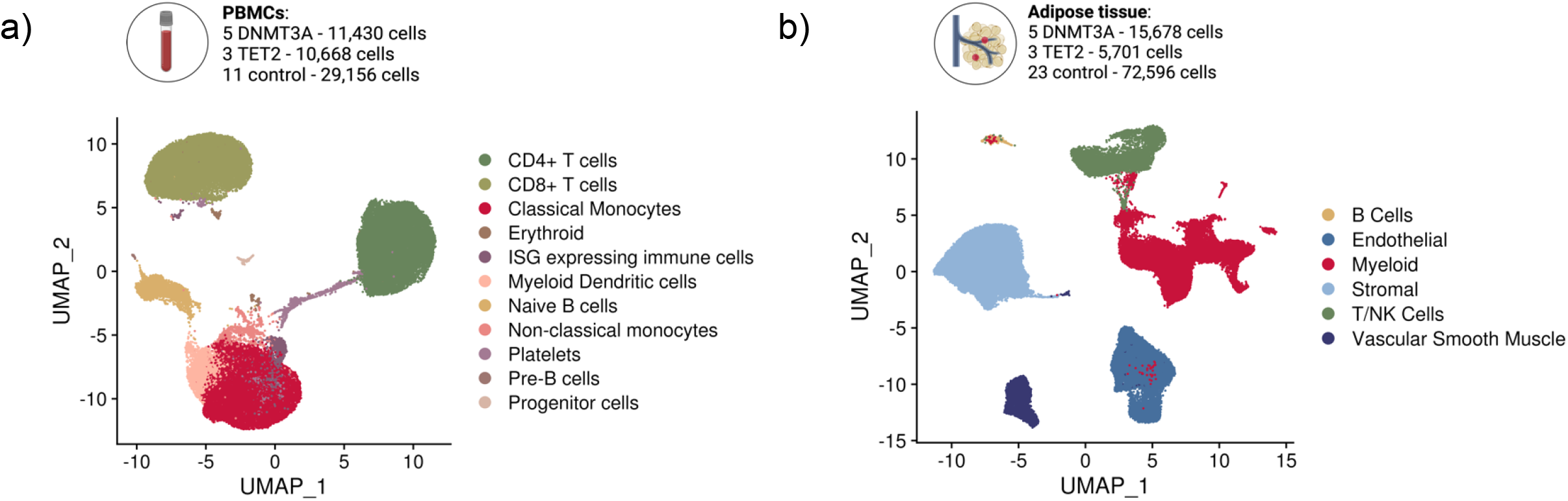
Single-cell RNA sequencing captures expected cell types in peripheral blood and adipose tissue. a) UMAP of peripheral blood mononuclear cells (PBMCs) from patients with TET2 and DNMT3A CHIP and controls. b) UMAP of adipose tissue cells from patients with TET2 and DNMT3A CHIP and controls.

### Monocytes with TET2 and DNMT3A mutations have decreased movement over endothelial cells

As CHIP mutations in the blood preferentially differentiate into monocytes, we next focused on characterizing the interaction between monocytes and endothelial cells in CHIP in the setting of diabetes. We used CellChat to predict the likelihood of interactions between monocytes and adipose tissue endothelial cells from scRNAseq data from our human patient cohort based on the expression of ligand-receptor pairs. We found several mutation-specific differences in signaling related to monocyte transendothelial migration^31–36^ (**Figure 2a, Figure S6, Table S1-2**). Notably, ITGB2 and ICAM signaling were significantly increased in DNMT3A CHIP compared to controls (p-value < 0.05) while CXCL signaling was significantly increased in TET2 CHIP compared to controls (p-value < 0.05).

**Figure 2:**
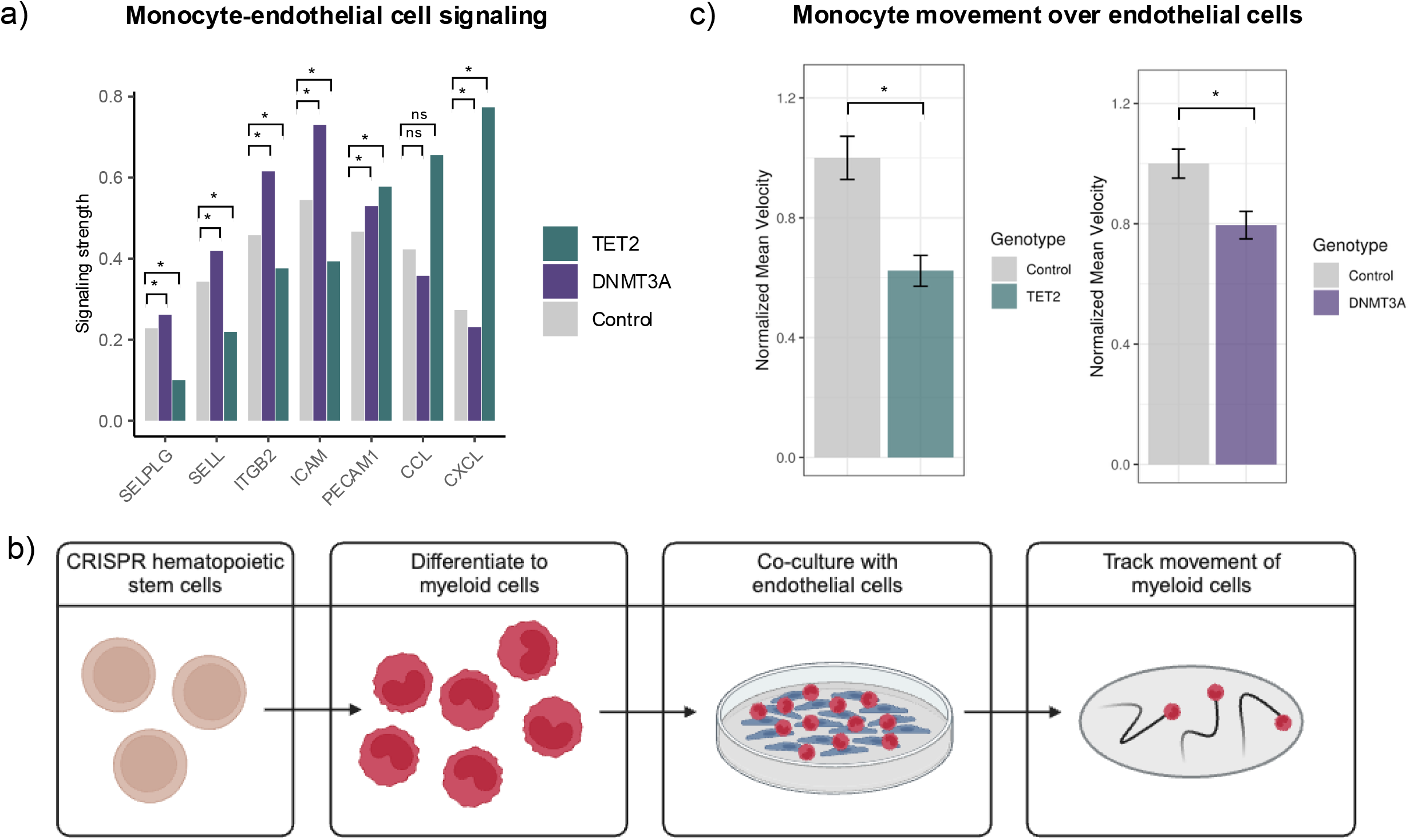
CHIP affects monocyte movement through alterations to monocyte-endothelial cell interactions. a) Barplot representing signaling strength between classical monocytes and endothelial cells for interactions related to transendothelial migration. Statistical significance evaluated with a Wilcoxon signed-rank test. * indicates p-value < 0.05. b) Schematic explaining *in vitro* model of monocyte movement over endothelial cells. c) Barplot showing normalized movement of monocytes with CHIP mutations and control monocytes over endothelial cells. Endothelial cells were cultured with dimethyl sulfoxide (DMSO) for 24 hours prior to co-culturing. Mean velocity for individual cells was normalized by dividing by the average velocity of control monocytes. Statistical significance was evaluated with a two-sample T test. * indicates p-value < 0.05.

We hypothesized that these differences in monocyte-endothelial signaling might be associated with differential physical interaction. We designed an *in vitro* experiment to quantify differences in the velocity of monocyte movement over a layer of vascular endothelial cells for monocytes carrying CHIP mutations, compared to controls (**Figure 2b**). We found that monocytes with CHIP mutations moved across endothelial cells at significantly lower velocity compared to controls for both *TET2*-(p-value = 2.6×10^-5^) and *DNMT3A*-mutated (p-value = 2.2 ×10^-3^) monocytes (**Figure 2c, Table S3-4**).

### In silico drug screen identifies drug perturbations that restore TET2 CHIP monocyte-endothelial interactions

We sought to restore normal interactions between CHIP monocytes and endothelial cells. To prioritize candidates for TET2 CHIP, we turned to the single-cell foundation model Geneformer.^28^ With contextual knowledge from pretraining on scRNAseq data from 30 million human cells, Geneformer prioritizes genes that play key roles in determining cell state. We found that CD14+ monocytes from patients with TET2 CHIP and controls were indistinguishable using traditional embeddings based on RNA (**Figure 3a**), but that Geneformer was able to clearly separate the two groups (**Figure 3b, Figure S1**). We then performed *in silico* perturbations and used Geneformer to predict changes to cell state. Geneformer suggested 11 druggable genetic targets likely to shift TET2 CHIP monocytes towards a control state (**Table S5**).

**Figure 3:**
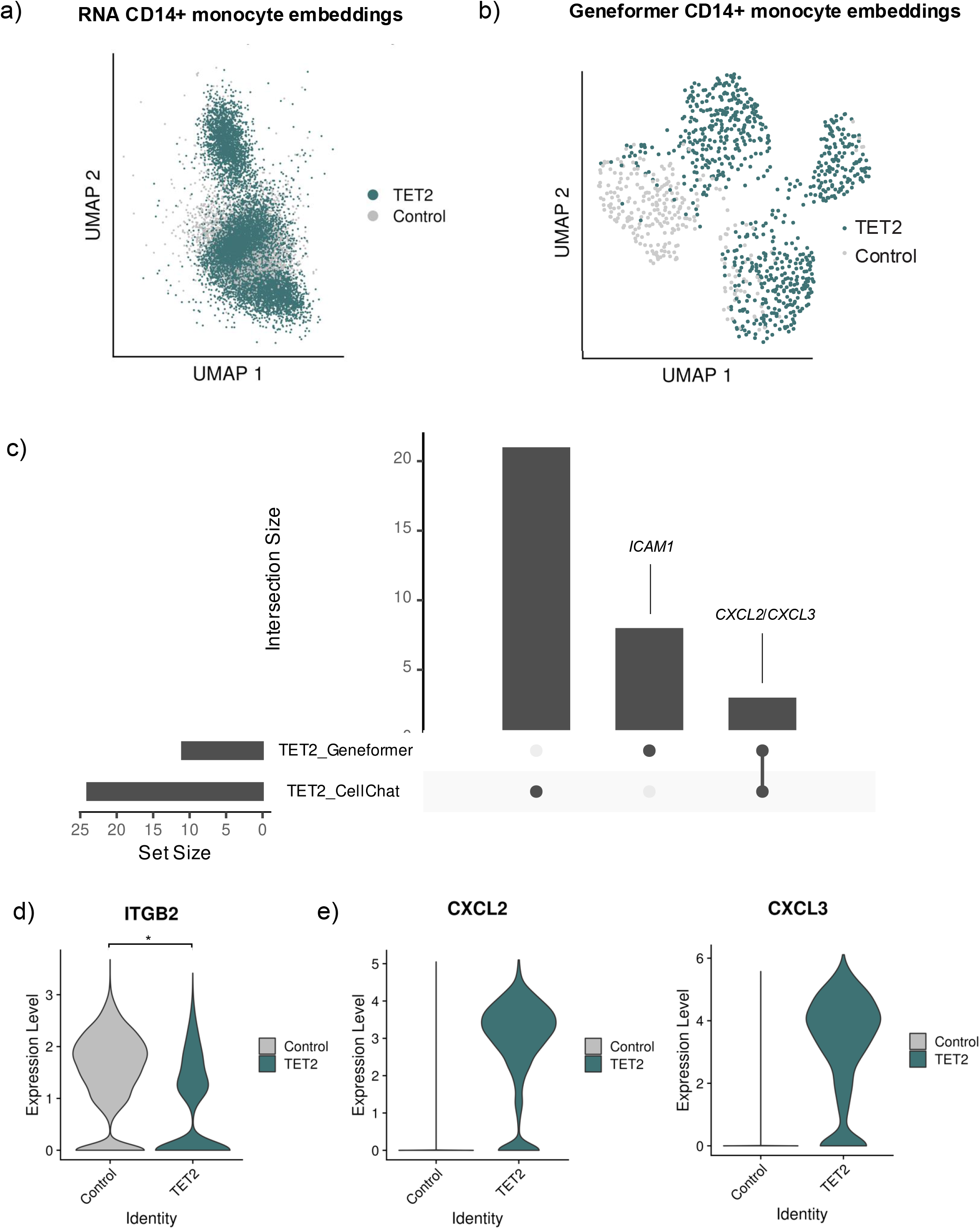
Single-cell foundation model nominates genetic perturbations capable of rescuing interactions between monocytes and endothelial cells in TET2 CHIP. a) UMAP of RNA-based embeddings representing CD14+ monocytes from patients with TET2 CHIP and controls. b) UMAP of Geneformer embeddings after fine-tuning to separate CD14+ monocytes from patients with TET2 CHIP and controls. c) Upset plot highlighting overlap of perturbation candidates identified by CellChat and Geneformer. d) Violin plot showing expression of ITGB2 in CD14+ monocytes. Statistical significance evaluated with a Wald test in a negative binomial generalized linear model (GLM). * indicates adjusted p-value < 0.05. e) Violin plots showing expression of CXCL2 and CXCL3 in CD14+ monocytes.

We compared these predictions to the ligand-receptor pairs identified by CellChat (**Figure 3c**). The two orthogonal approaches nominated two ligand-receptor pair candidates: *ITGB2-ICAM1* and *CXCL2*/*CXCL3-CXCR2*. While expression of *ITGB2* was lower in TET2 monocytes than controls (**Figure 3d**), *in silico* knockout of its binding partner, *ICAM1* was predicted to shift both TET2 monocytes to a control state. Expression of *CXCL2* and *CXCL3* were higher in TET2 monocytes compared to controls (**Figure 3e**), but expression level did not meet the threshold for evaluation with differential expression. *In silico* perturbations from Geneformer, however, predicted that inhibition of *CXCL2* and *CXCL3* would restore the transcriptional profile of TET2 CHIP monocytes to that of control monocytes.

We tested the effect of inhibiting these interactions on monocyte movement over endothelial cells *in vitro*. We chose to target *ICAM1*, which is expressed on endothelial cells, with lifitegrast sodium (SAR1118). Incubation of endothelial cells with this *ICAM1* inhibitor for 24 hours before co-culturing with monocytes increased monocyte movement among *TET2*-mutated monocytes at a concentration of 2nM, compared to *TET2*-mutated monocytes cultured with DMSO (**Figure 4a, Table S1**; p-value = 1.2×10^-2^). *TET2*-mutated monocyte velocity was significantly less than control monocyte velocity when both were treated with the same drug and concentration for DMSO (p-value = 2.6×10^-5^), 0.1nM SAR1118 (p-value = 1.7×10^-5^), and 1nM SAR1118 (p-value = 2.6×10^-2^), but when treated with 2nM SAR1118, there was not a statistically significant difference in velocity between *TET2*-mutated monocytes and control monocytes (p-value = 0.83), suggesting that the effect of the drug treatment has specificity for *TET2*-mutated monocytes.

**Figure 4:**
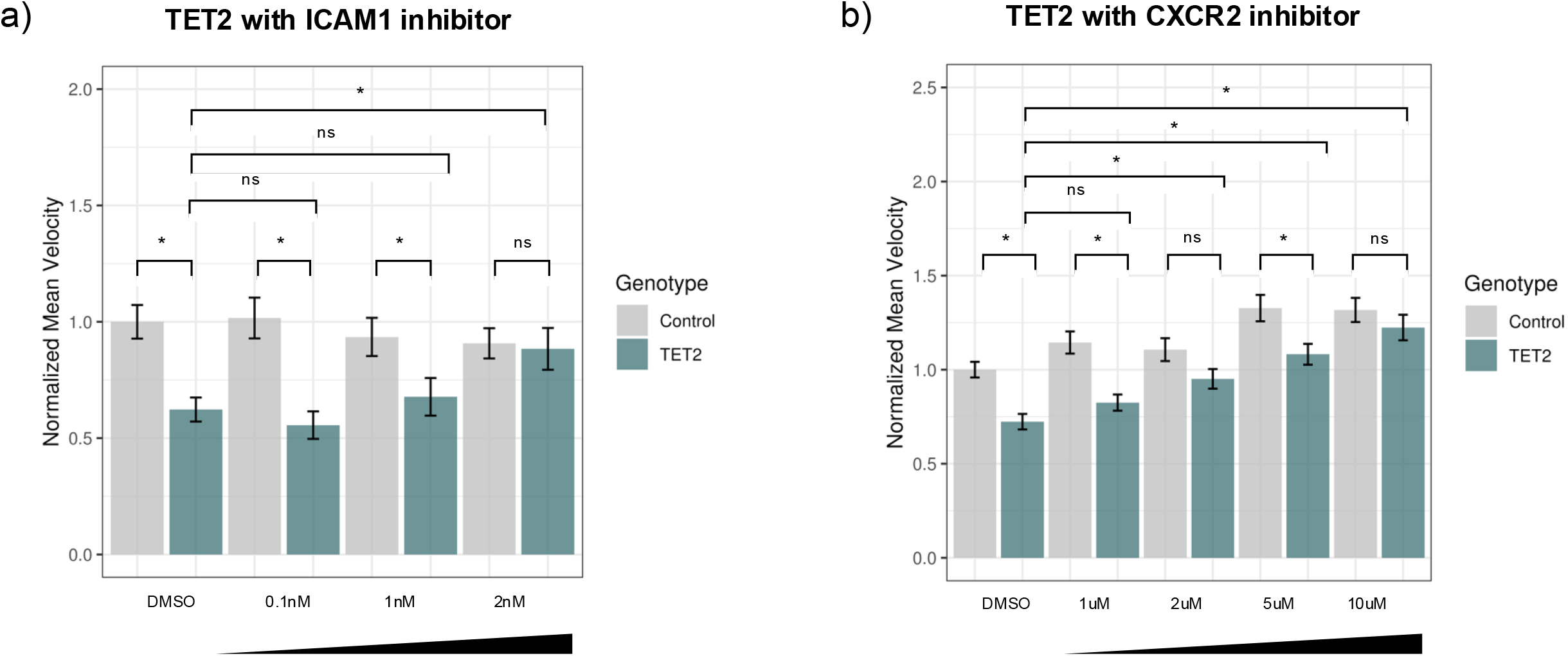
ICAM and CXCR2 are effective targets for restoring movement of TET2-mutated monocytes over endothelial cells. a) Barplot showing normalized mean velocity of *TET2*-mutated monocytes over endothelial cells that were cultured with DMSO or with SAR1118, an ICAM1 inhibitor, at 0.1nM, 1nM, and 2nM for 24 hours. Statistical significance evaluated with a two-sample T test. * indicates p-value < 0.05. b) Barplot showing normalized mean velocity of *TET2*-mutated monocytes over endothelial cells after monocytes were cultured for 24 hours with DMSO or AZD5069, a CXCR2 inhibitor, at 1uM, 2uM, 5uM, and 10uM.

We also chose to target *CXCR2*, which is expressed on monocytes, with AZD5069. Incubation of monocytes with this *CXCR2* inhibitor for 24 hours before co-culturing with endothelial cells increased monocyte velocity for *TET2*-mutated monocytes, at a concentration of 2uM, 5uM, and 10uM, compared to *TET2*-mutated monocytes cultured with DMSO (**Figure 4b, Table S1**; p-value = 6.5×10^-4^, 2.6×10^-7^, 4.7×10^-10^). *TET2*-mutated monocyte velocity was significantly less than control monocyte velocity when both were treated with the same drug and concentration for DMSO (p-value = 2.8×10^-6^), 1uM AZD5069 (p-value = 1.4×10^-5^), and 5uM AZD5069 (p-value = 6.1×10^-3^), but when treated with 2uM AZD5069 (p-value = 5.3×10^-2^) and 10uM AZD5069 (p-value = 0.32), there was not a statistically significant difference in velocity between *TET2*-mutated monocytes and control monocytes, again suggesting that the effect of the drug treatment has specificity for *TET2*-mutated monocytes.

## Discussion

Here, we conducted the first multi-tissue scRNAseq analysis of CHIP in humans, using samples from peripheral blood and adipose tissue from patients with diabetes. We identified increased likelihood of interactions between monocytes and endothelial cells in both TET2 and DNMT3A CHIP, based on expression of ligand-receptor pairs. We demonstrated that monocytes carrying CHIP mutations move over endothelial cells at a decreased velocity, compared to monocytes without CHIP mutations. We employed a single-cell foundation model to perform a genomewide *in silico* perturbation screen and identified pharmacologic perturbations capable of attenuating the effects of *TET2* mutations on monocyte-endothelial cell interactions. These findings permit several conclusions.

First, we demonstrated that interactions of monocytes and endothelial cells are altered in CHIP in a mutation-specific manner. Previous work has investigated the interaction of CHIP-mutated monocytes and endothelial cells in mouse models and with cancer cell lines,^37,38^ but this is the first study to demonstrate such effects in human patient samples. Furthermore, prior work has been limited to *DNMT3A*-mutated cells. Here, we show that mutations in *TET2* also alter monocyte-endothelial interactions, affecting the velocity of monocyte movement over endothelial cells. Decreased velocity of monocytes over endothelial cells may increase the likelihood of CHIP-mutated monocytes crossing the endothelial barrier. We find that while both genotypes decrease the velocity of monocyte movement over endothelial cells, the mechanism by which adhesion increases may be mutation-specific. We found increased *ITGB2*-*ICAM1* signaling in DNMT3A CHIP and increased *CXCL2*/*CXCL3*-*CXCR2* signaling in TET2 CHIP. These differences may explain variability in vascular disease risk observed in TET2 and DNMT3A CHIP.^3,4^

Second, for the first time we show that single-cell foundation models are capable of identifying pharmacological targets that rescue an *in vitro* phenotype. As predicted by the single-cell foundation model Geneformer, we found that inhibition of ICAM1 in endothelial cells and CXCR2 in monocytes rescued the monocyte movement over endothelial cells in an *in vitro* model of TET2 CHIP. While traditional scRNAseq analysis methods did not converge on *ICAM1* and *CXCR2* for TET2 CHIP, Geneformer suggested that both treatments would be effective. Through *in vitro* experimentation, we found that both drugs increased the velocity of *TET2*-mutated monocytes over endothelial cells. These findings are relevant for the development of mutation-specific treatments to address the increased risk of CHIP-mediated cardiovascular disease. Notably, this set of experiments represents, to our knowledge, the first use of an AI foundation model to identify and experimentally test a vascular disease target. Therefore, this study provides an early example of what we expect will be a paradigm shift accelerating the rate of therapeutic discovery for cardiovascular medicine.

Overall, through integrative single cell analysis of peripheral blood and adipose tissue, *in silico* perturbations, and *in vitro* validation experiments, we identified mechanisms by which mutated blood cells may contribute to vascular disease risk among patients with CHIP.

## Acknowledgements

This study was funded by the following: K23DK135414 (SSB, JRK), Doris Duke CSDA 2021193 (CNW), K23 HL156759 (CNW), Burroughs Wellcome Fund 1021480 (CNW), R01 DK112262 (JRK), the Tennessee Center for AIDS Research grant P30 AI110527 (Supplement CNW, JK), the Vanderbilt Flow Cytometry Shared Resource is supported by the Vanderbilt Ingram Cancer Center (P30 CA068485). This work was also supported by NIH grants DP5 OD029586, R01 AG088657, R01 AG083736, a Burroughs Wellcome Fund Career Award for Medical Scientists, the E.P. Evans Foundation, a Pew-Stewart Scholar for Cancer Research award, supported by the Pew Charitable Trusts and the Alexander and Margaret Stewart Trust, a Hevolution/AFAR New Investigator Award in Aging Biology and Geroscience Research (AGB). ACP and JCV have been supported by the NIH training grant T32 GM145734-01. YP and HKG have been supported by the NIH training grant T32 GM007347.

Imaging was performed through the use of the Vanderbilt Cell Imaging Shared Resource (supported by NIH grants CA68485, DK20593, DK58404, DK59637, EY08126, S10 OD021630, and S10 RR027396).

## Disclosures

None.

## Graphical abstract

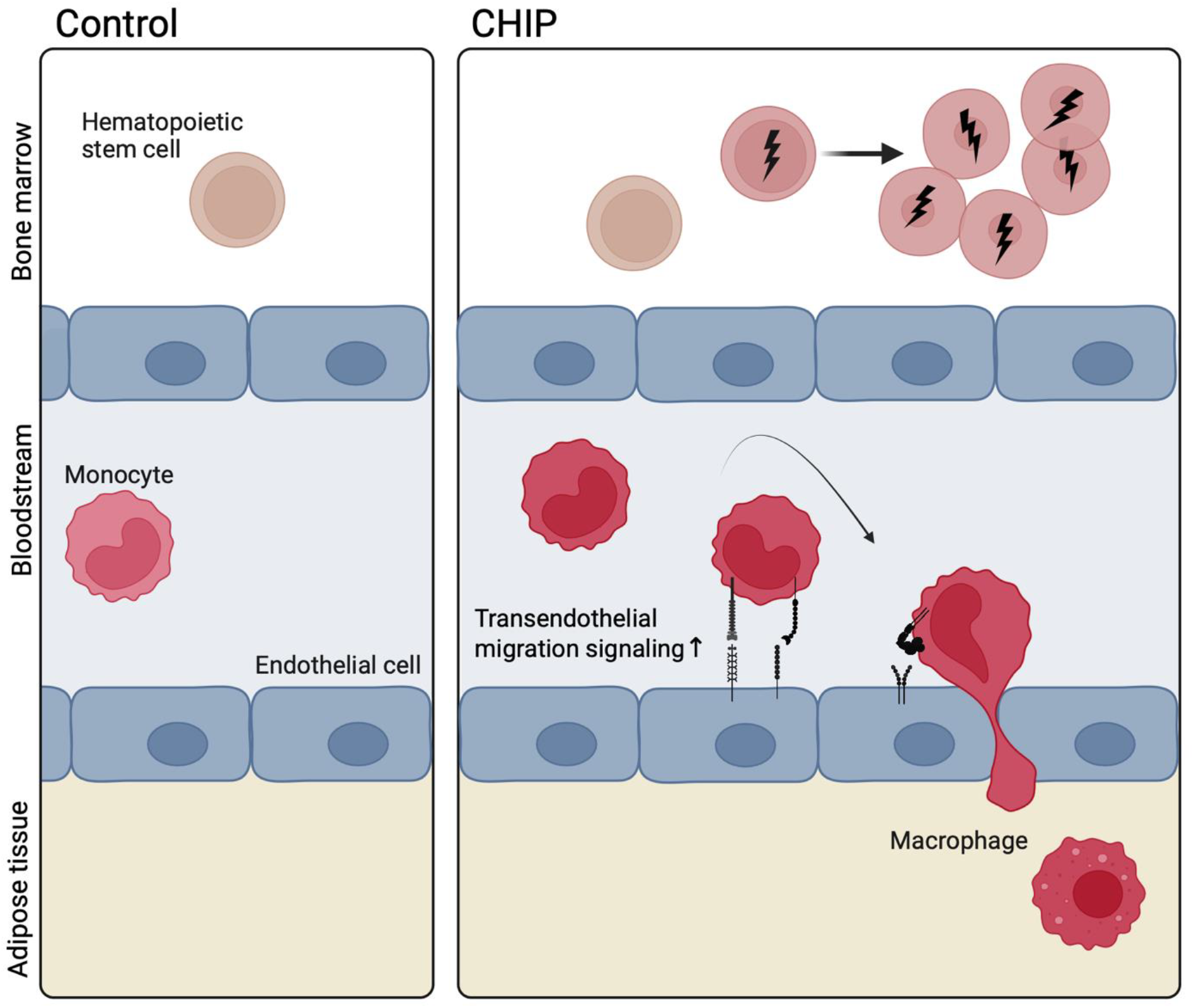

## Notes

### Clinical Trial

NCT04451980

### Author Declarations

The study was approved by the Vanderbilt University Medical Center Institutional Review Board (identifiers: 210022 and 201583).

### Summary of Updates

Cohort size was increased, in vitro validation of monocyte-endothelial cell interaction was performed, and targets to restore normal interactions were identified with an in silico genomewide perturbation screen.

